# Is the infection of the SARS-CoV-2 Delta variant associated with the outcomes of COVID-19 patients?

**DOI:** 10.1101/2021.10.05.21262783

**Authors:** Gunadi, Mohamad Saifudin Hakim, Hendra Wibawa, Marcellus, Vivi Setiawaty, Slamet, Ika Trisnawati, Endah Supriyati, Riat El Khair, Kristy Iskandar, Afiahayati, Siswanto, Irene, Nungki Anggorowati, Edwin Widyanto Daniwijaya, Dwi Aris Agung Nugrahaningsih, Yunika Puspadewi, Dyah Ayu Puspitarani, Irene Tania, Khanza Adzkia Vujira, Muhammad Buston Ardlyamustaqim, Gita Christy Gabriela, Laudria Stella Eryvinka, Bunga Citta Nirmala, Esensi Tarian Geometri, Abirafdi Amajida Darutama, Anisa Adityarini Kuswandani, Lestari, Sri Handayani Irianingsih, Siti Khoiriyah, Ina Lestari, Nur Rahmi Ananda, Eggi Arguni, Titik Nuryastuti, Tri Wibawa, on behalf of the Yogyakarta-Central Java COVID-19 study group

## Abstract

**Background:** SARS-CoV-2 Delta variant (B.1.617.2) has been responsible for the current increase in COVID-19 infectivity rate worldwide. We compared the impact of the Delta variant and non-Delta variant on the COVID-19 outcomes in patients from Yogyakarta and Central Java provinces, Indonesia.

**Methods:** We ascertained 161 patients, 69 with the Delta variant and 92 with the non-Delta variant. The Illumina MiSeq next-generation sequencer was used to perform the whole genome sequences of SARS-CoV-2.

**Results:** The mean age of patients with Delta and the non-Delta variant was 27.3 ± 20.0 and 43.0 ± 20.9 (*p*=3×10^−6^). The patients with Delta variant consisted of 23 males and 46 females, while the patients with the non-Delta variant involved 56 males and 36 females (*p*=0.001). The Ct value of the Delta variant (18.4 ± 2.9) was significantly lower than the non-Delta variant (19.5 **±** 3.8) (*p*=0.043). There was no significant difference in the hospitalization and mortality of patients with Delta and non-Delta variants (*p*=0.80 and 0.29, respectively). None of the prognostic factors was associated with the hospitalization, except diabetes with an OR of 3.6 (95% CI=1.02-12.5; *p*=0.036). Moreover, the patients with the following factors have been associated with higher mortality rate than patients without the factors: age ≥65 years, obesity, diabetes, hypertension, and cardiovascular disease with the OR of 11 (95% CI=3.4-36; *p*=8×10^−5^), 27 (95% CI=6.1-118; *p*=1×10^−5^), 15.6 (95% CI=5.3-46; *p*=6×10^−7^), 12 (95% CI=4-35.3; *p*=1.2×10^−5^), and 6.8 (95% CI=2.1-22.1; *p*=0.003), respectively. Multivariate analysis showed that age ≥65 years, obesity, diabetes, and hypertension were the strong prognostic factors for the mortality of COVID-19 patients with the OR of 3.6 (95% CI=0.58-21.9; *p*=0.028), 16.6 (95% CI=2.5-107.1; *p*=0.003), 5.5 (95% CI=1.3-23.7; *p*=0.021), and 5.8 (95% CI=1.02-32.8; *p*=0.047), respectively.

**Conclusions:** We show that the patients infected by the SARS-CoV-2 Delta variant have a lower Ct value than the patients infected by the non-Delta variant, implying that the Delta variant has a higher viral load, which might cause a more transmissible virus among humans. However, the Delta variant does not affect the COVID-19 outcomes in our patients. Our study also confirms the older age and comorbidity increase the mortality rate of COVID-19 patients.

## Introduction

The SARS-CoV-2 variants of concern (VOC), including Alpha, Beta, Gamma, and Delta, has attracted public health authorities due to its capability on higher transmission, the possibility of affecting COVID-19 severity, and the impact of the effectiveness of public health measures, diagnosis, treatment, and vaccines [1,2,3]. SARS-CoV-2 Delta variant (B.1.617.2) has been responsible for the current increase in COVID-19 infectivity rate worldwide [4, 5, 6,7].

However, our knowledge about the role of the Delta variant on the COVID-19 outcomes is still very limited [8, 9]. Moreover, several prognostic factors have been associated with COVID-19 illness [10,11,12]. Here, we compared the impact of the Delta variant and non-Delta variant on the COVID-19 outcomes in patients from Yogyakarta and Central Java provinces, Indonesia.

## Material and Methods

### Patients

We ascertained 161 COVID-19 patients from Yogyakarta and Central Java provinces, 79 males and 82 females. The outcomes of COVID-19 patients were hospitalization and mortality. The Ethics Committee the Faculty of Medicine, Public Health and Nursing, Universitas Gadjah Mada/Dr. Sardjito Hospital approved our study (KE/FK/0563/EC/2020).

### Prognostic Factors

According to previous studies, we associated the following prognostic factors with the hospitalization and mortality of COVID-19 patients: sex, age, comorbidity, and smoking [10,11,12].

### SARS-CoV-2 whole-genome sequencing

First, Single-stranded cDNA was synthesized from RNA extracted from the viral transport medium of patients with COVID-19 using SuperScript™ III First-Strand Synthesis System (Thermo Fisher Scientific, MA, United States). Then, the second strand was synthesized using COVID-19 ARTIC v3 primer pool design by SARS-CoV-2 ARTIC Network using Phusion™ High-Fidelity DNA Polymerase (Thermo Fisher Scientific, MA, United States). The library preparations were performed using the Illumina DNA Prep (Illumina, California, United States). The Illumina MiSeq next-generation sequencer was used to perform the whole genome sequences of SARS-CoV-2. The assembly of our sample genomes was mapped into the reference genome from Wuhan, China (hCoV-19/Wuhan/Hu-1/2019, GenBank accession number: NC_045512.2) using Burrow-Wheeler Aligner (BWA) algorithm embedded in UGENE v. 1.30 [13].

### Phylogenetic study

We used a dataset of 250 available SARS-CoV-2 genomes extracted from GISAID from our region and others (Acknowledgment Table is provided in Supp. Table 1) to reconstruct the phylogenetic tree. Multiple nucleotide sequence alignment was performed using the MAFFT program (https://mafft.cbrc.jp/alignment/server/). We used the neighbor-joining statistical method with 1,000 bootstrap replications [14,15] to construct a phylogenetic tree from 29.420 nucleotide length of the open reading frame (ORF) of SARS-CoV-2, followed by computation of the evolutionary distances and model the rate variation among sites by the Kimura 2-parameter method and the gamma distribution with estimated shape parameter (α) for the dataset, respectively [16]. We used the DAMBE version 7 [17] to calculate the estimation of the α gamma distribution, MEGA version 10 (MEGA X) [18] for phylogenetic reconstruction, and FigTree to visualize the Newick tree output from MEGA X.

### Statistical analysis

We presented data as mean ± SD and frequency (percentage). Chi-square or Fisher exact tests with 95% confidence interval (CI) were used to find any significant association between independent variables and COVID-19 outcomes. A logistic regression test was used for multivariate analysis. The *p*-value of <0.05 was considered significant.

## Results

### Phylogenetic analysis

Phylogenetic analysis showed that about 42.86% of SARS-CoV-2 collected from Central Java and Yogyakarta provinces belonged to B.1.617.2 lineage (Delta variant), while 57.14% clustered in 14 different lineages based on the Pango nomenclature (B, B.1, B.1.1, B.1.459, B.1.456, B.1.459, B.1.462, B.1.466.2, B.1.468, B.1.470, B.1.570, B.1.1.236, B.1.36.19, B.1.1.398) and seven virus samples were not belonging to any of Pango lineages. They are called “none” (Fig.1). Except for Delta variants, none of the virus samples collected from our study belonged to any VOC or variant of interest (VOI) according to WHO labels for naming SARS-CoV-2 variants. However, we found that about 13.04% of virus samples from Central Java and Yogyakarta provinces were clustered into B.1.466.2 lineage (Fig.1) that is currently designated by WHO as a variant of alert for further monitoring.

**Figure 1.**
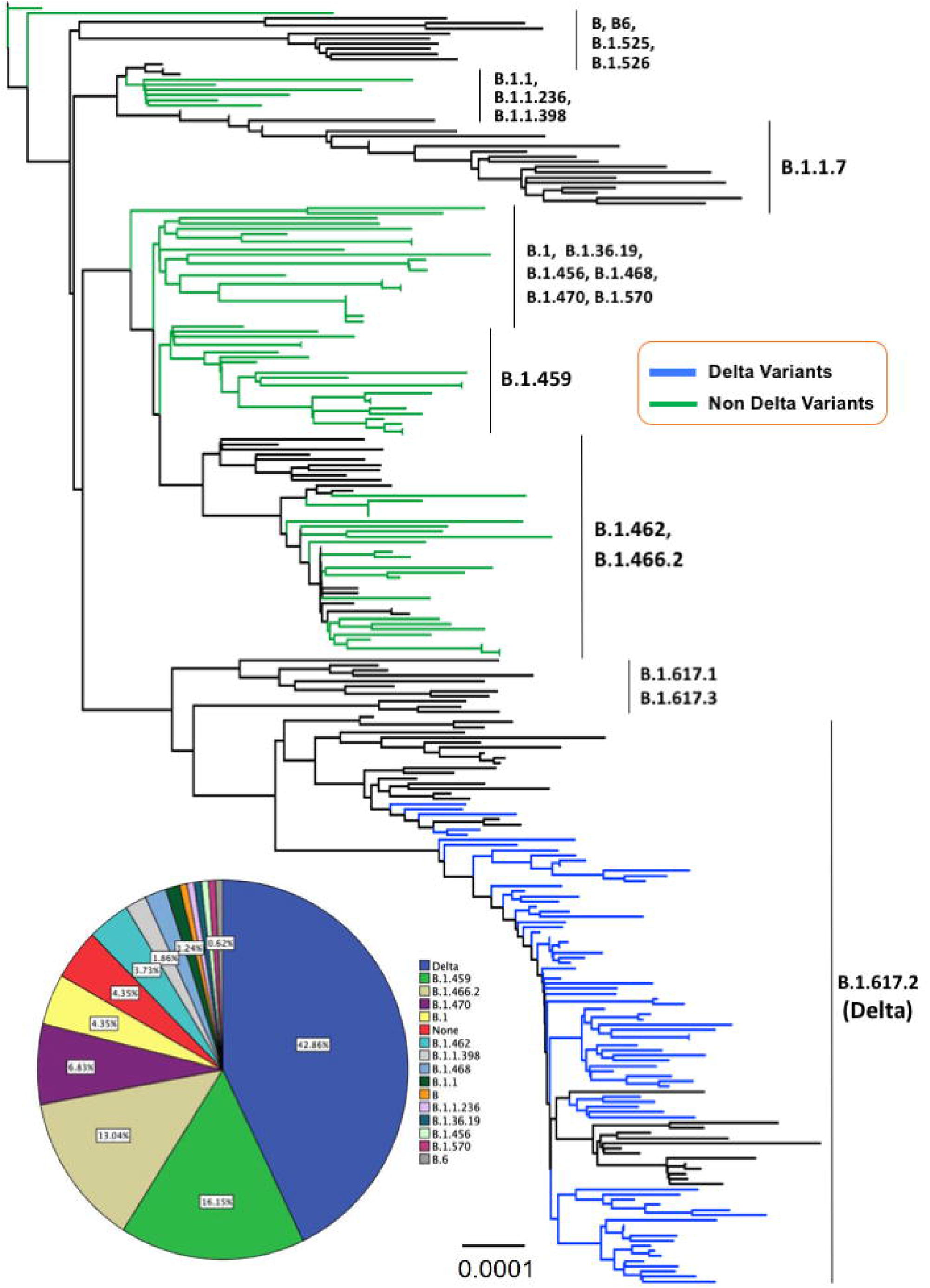
(a) The evolutionary history was inferred using the Neighbor-Joining method conducted in MEGA-X. The evolutionary distances were computed using the Kimura 2-parameter method with 1000 bootstrap replication and are in the units of the number of base substitutions per site (0.0001) shown in the bottom tree. This analysis involved 250 nucleotide sequences with a total of 29.420 positions in the final dataset and all ambiguous positions were removed for each sequence pair (pairwise deletion option). The Delta variant taxa are indicated in blue line, whereas non-Delta variant taxa appeared in green line. (b) A pie chart illustrates the proportion of Delta variant and non-Delta samples detected in the present study.

### Characteristics of COVID-19 patients

The mean age of patients with Delta and the non-Delta variant was 27.3 ± 20.0 and 43.0 ± 20.9 (*p*=3×10^−6^). The patients with the Delta variant consisted of 23 males and 46 females, while the patients with the non-Delta variant involved 56 males and 36 females (*p*=0.001). The Ct value of the Delta variant (18.4 **±** 2.9) was significantly lower than the non-Delta variant (19.5 **±** 3.8) (*p*=0.043 (Table 1).

**Table 1.** Characteristics of COVID-19 patients from Yogyakarta and Central Java provinces, Indonesia.

| Characteristics | Total (N=161)<br>N (%); mean ± SD | Delta variant<br>(N=69) N (%);<br>mean ± SD | Non-Delta variant<br>(N=92) N (%);<br>mean ± SD | p-value |
| --- | --- | --- | --- | --- |
| Ct value | $19.1 \pm 3.5$ | $18.4 \pm 2.9$ | $19.5 \pm 3.8$ | 0.043* |
| Age (years) | $36.3 \pm 21.9$ | $27.3 \pm 20.0$ | $43.0 \pm 20.9$ | $3 \times 10^{-6}$* |
| ▪ $\geq 65$ | 17 (10.6) | 1 (1.4) | 16 (17.4) | $1 \times 10^{-6}$* |
| ▪ $< 65$ | 98 (60.9) | 35 (50.7) | 63 (68.5) | |
| ▪ $< 18$ | 46 (28.6) | 33 (47.8) | 13 (14.1) | |
| Sex |  |  |  |  |
| ▪ Male | 79 (49.1) | 23 (33.3) | 56 (60.9) | 0.001* |
| ▪ Female | 82 (50.9) | 46 (66.7) | 36 (39.1) |  |
| Comorbidities |  |  |  |  |
| ▪ Obesity | 10 (6.2) | 3 (4.3) | 7 (7.6) | 0.51 |
| ▪ Diabetes | 26 (16.1) | 7 (10.1) | 19 (20.7) | 0.07 |
| ▪ Hypertension | 22 (13.7) | 4 (5.8) | 18 (19.6) | 0.012* |
| ▪ Cardiovascular disease | 15 (9.3) | 4 (5.8) | 11 (12) | 0.18 |
| ▪ Chronic kidney disease | 2 (1.2) | 0 | 2 (2.2) | 0.50 |
| Smoking | 5 (3.1) | 1 (1.4) | 4 (4.3) | 0.39 |

\*, significant ( $p < 0.05$ ); SD, standard deviation

### Association between independent variables and COVID-19 patients’ outcomes

Next, we associated the independent variables with the COVID-19 outcomes, hospitalization, and mortality. There were no significant differences in the hospitalization and mortality of patients with Delta and non-Delta variants (*p*=0.80 and 0.29, respectively) (Table 2). None of the prognostic factors was associated with the hospitalization, except comorbidity of diabetes with the OR of 3.6 (95% CI=1.02-12.5; *p*=0.036) (Table 2). Moreover, the patients with the following factors have been associated with higher mortality rate than patients without these factors: age ≥65 years, obesity, diabetes, hypertension, and cardiovascular disease with OR of 11 (95% CI=3.4-36; *p*=8×10^−5^), 27 (95% CI=6.1-118; *p*=1×10^−5^), 15.6 (95% CI=5.3-46; *p*=6×10^−7^), 12 (95% CI=4-35.3; *p*=1.2×10^−5^), and 6.8 (95% CI=2.1-22.1; *p*=0.003), respectively (Table 2).

**Table 2.**
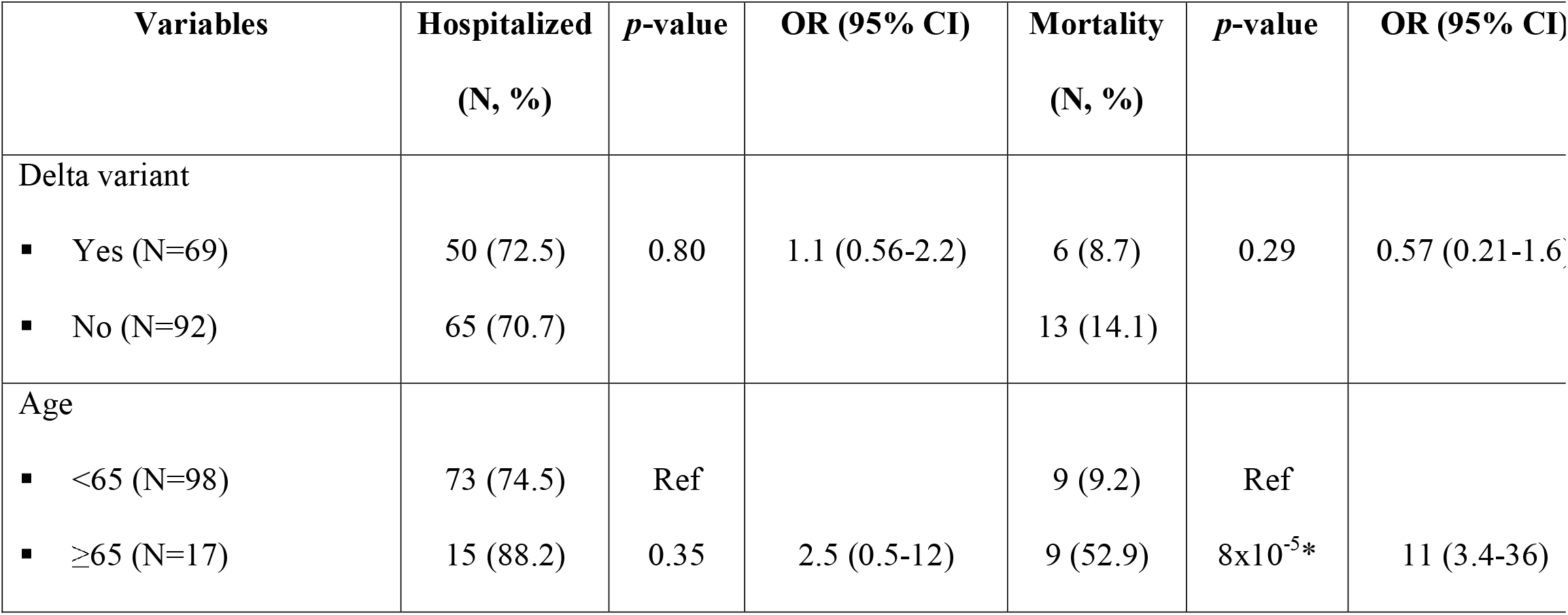
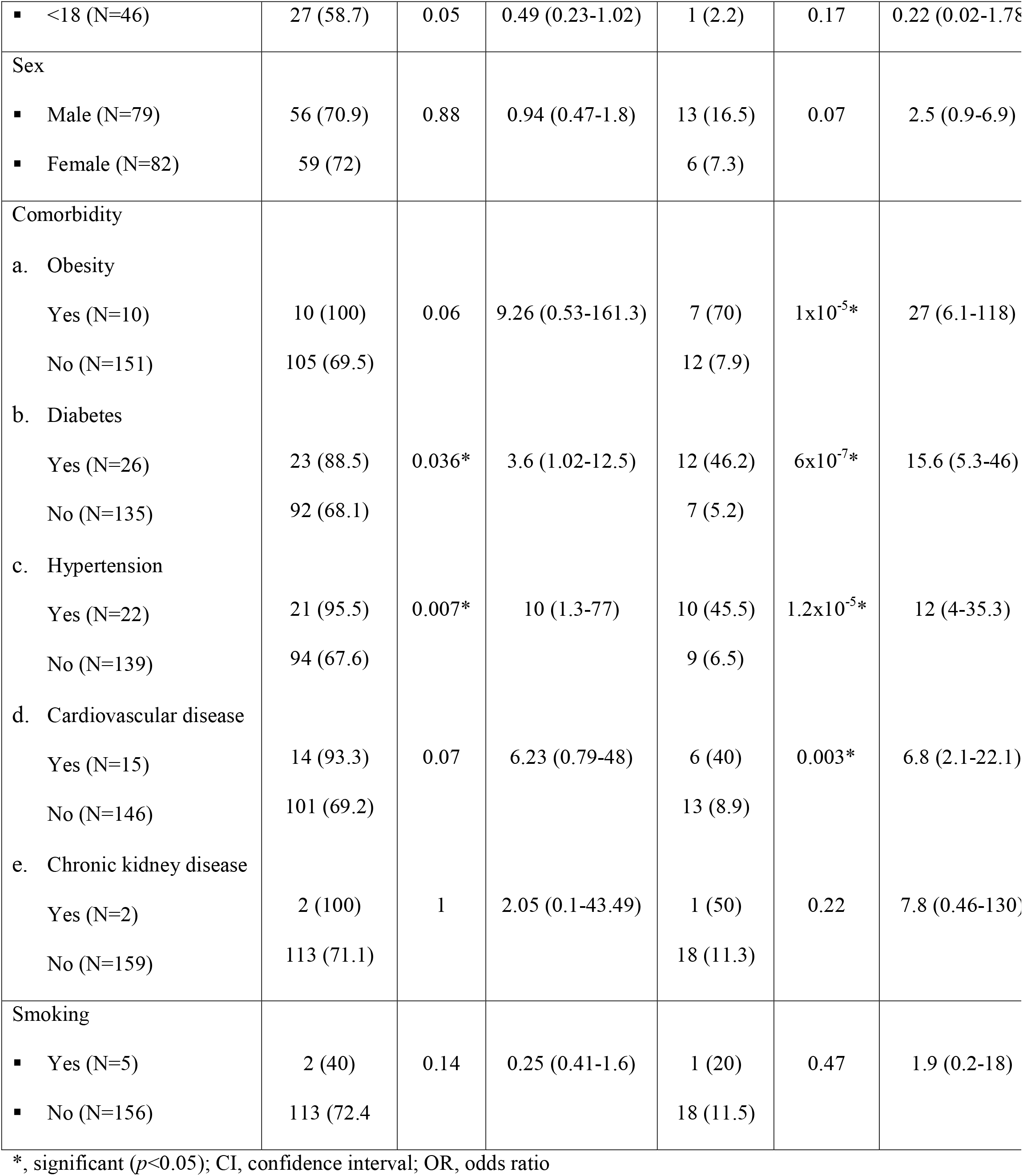
Association between independent variables and of COVID-19 patients’ outcomes.

### Multivariate analysis

Subsequently, we performed a multivariate analysis to find an independent factor affecting the COVID-19 outcomes. Multivariate analysis showed that age ≥65 years, obesity, diabetes, and hypertension were strong prognostic factors for the mortality of COVID-19 patients with the OR of 3.6 (95% CI=0.58-21.9; *p*=0.028), 16.6 (95% CI=2.5-107.1; *p*=0.003), 5.5 (95% CI=1.3-23.7; *p*=0.021), and 5.8 (95% CI=1.02-32.8; *p*=0.047), respectively. In addition, no prognostic factors were associated with the hospitalization of COVID-19 patients (Table 3).

**Table 3.** Multivariate analysis of the association between independent variables and COVID patients’ outcomes

| Variables | Hospitalized |  | Mortality |  |
| --- | --- | --- | --- | --- |
|  | OR (95% CI) | *p*-value | OR (95% CI) | *p*-value |
| Delta variant | 1.3 (0.6-2.8) | 0.47 | 3.6 (0.58-21.9) | 0.167 |
| Age ($\geq 65$ years) | 0.7 (0.1-5.1) | 0.79 | 11.5 (1.3-102.6) | 0.028* |
| Sex (male) | 0.9 (0.46-2.1) | 0.98 | 3.2 (0.65-16) | 0.15 |
| Comorbidity |  |  |  |  |
| ▪ Obesity | - | 0.99 | 16.6 (2.5-107.1) | 0.003* |
| ▪ Diabetes | 1.68 (0.41-6.8) | 0.46 | 5.5 (1.3-23.7) | 0.021* |
| ▪ Hypertension | 10.4 (0.92-118) | 0.058 | 5.8 (1.02-32.8) | 0.047* |
| ▪ Cardiovascular disease | 3 (0.3-28.8) | 0.32 | 0.7 (0.08- 6.2) | 0.75 |
| ▪ Chronic kidney disease | - | 0.99 | 1.2 (0.015-96.6) | 0.92 |
| Smoking | 0.11 (0.008-1.4) | 0.09 | 0.5 (0.003-74.4) | 0.78 |

\*, significant ( $p<0.05$ ); CI, confidence interval; OR, odds ratio

## Discussion

We are able to show that the Ct value of the patients infected by the SARS-CoV-2 Delta variant has a lower Ct value than the patients infected by the non-Delta variant. Our finding is compatible with previous studies [19,20; 21]. Our finding supports that the higher viral load of the Delta variant results in its characteristic of being more transmissible among humans [19,22]. It is associated with a higher reproductive number (R0) of the Delta variant (R0=7) than the parental SARS-CoV-2 and the Alpha variant [23]. In addition, the Delta variant also has a significantly longer duration (18 days) of Ct value ≤30 than the original strain (13 days) [20].

Again, this evidence implies higher transmissibility of the Delta variant than other strains.

We also show that the Delta variant is not associated with the mortality and hospitalization of COVID-19 patients. Our findings are different from previous reports [8,20,24]. Sheikh *et al*. [2021] showed that the risk of hospitalization is twice higher in patients with Delta variant than Alpha variant. Fisman *et al*. [2021] revealed that the risk of hospitalization, admission of ICU, and mortality is significantly higher in the Delta variant than in N501Y-positive variants. They suggested that the Delta variant is more virulent than previous VOCs [24]. Ong *et al*. [2021] showed that the patients with the Delta variant showed more severe COVID-19 than the original variant. These differences might be due to differences in the host’s genetic background [25]. They identified a novel susceptibility locus for severe COVID-19 at 3p21.31 gene cluster, consisting of *SLC6A20, LZTFL1, CCR9, FYCO1, CXCR6*, and *XCR1*, that their functions are related to COVID-19 [25]. Further study is necessary to identify the genetic susceptibility locus for severe COVID-19 in our patients. Another difference between our study and previous reports is that we compared the outcomes of the Delta variant and non-VOC, while other reports compared the Delta variant and other VOCs [8; 24].

Interestingly, approximately 50% of Delta variant infected children, higher than non-Delta variant (∼15%). Similar findings were reported by a previous study showing the frequency of Delta variant is higher in aged 5-9 years than non-Delta variant [8]. While the Delta variant infected more females than males, in contrast, the non-Delta variant infected more males than females. However, multivariate analysis did not show an association between sex and COVID-19 outcomes. It is similar to a study by Ong *et al*. [2021]. It should be noted that the impact of sex on the COVID-19 outcomes is still controversial [10,20, 26,27]. While some studies showed that male has a higher risk for severe COVID-19 [10,26], other reports do not support the association [20, 27].

Our findings reveal the older age and comorbidities, including obesity, diabetes, and hypertension, are independent prognostic factors for the mortality of COVID-19 patients. Our findings were similar to previous studies [10, 20,26]. Several mechanisms have been proposed for the increased risk of COVID-19 in diabetes patients, including an elevated level of ACE-2 receptors and furin, and dysregulated immune response, while the following factors contribute for the obesity to be associated with the worse prognosis of COVID-19: the compromised ventilation at the base of the lung and immune response [28]. The use of the antihypertension drug, particularly ACE-2 inhibitors and angiotensin receptor blockers, is associated with the upregulated expression of the ACE-2 receptor, resulting in a higher possibility of respiratory failure [28]. Unfortunately, we do not have complete data on the use of ACE-2 inhibitors and angiotensin receptor blockers in our patients. Therefore, it is challenging to conclude that the increased mortality of COVID-19 in hypertension patients is due to antihypertension.

Our retrospective design and small sample size were the weakness of our study. Moreover, we only determined the impact of Delta variant and some comorbidities on the COVID-19 outcomes by overall means without considering other variables affecting the data, including other mutations in the SARS-CoV-2 genome and vaccination history.

## Conclusions

We show that the patients infected by the SARS-CoV-2 Delta variant have a lower Ct value than the patients infected by the non-Delta variant, implying that the Delta variant has a higher viral load, which might cause a more transmissible virus among humans. However, the Delta variant does not affect the COVID-19 outcomes in our patients. Our study also confirms that older age and comorbidity increase the mortality rate of COVID-19 patients.

## Data Availability

All data generated or analyzed during this study are included in the submission. The sequence and metadata are shared through GISAID (www.gisaid.org).

## List of abbreviations

CI: confidence interval
OR: odds ratio
VOC: variant of concern

## Declarations

### Ethics approval and consent to participate

This study was approved by the Medical and Health Research Ethics Committee, Faculty of Medicine, Public Health and Nursing, Universitas Gadjah Mada/Dr. Sardjito Hospital, Yogyakarta, Indonesia (KE/FK/0563/EC/2020). The research has been performed following the Declaration of Helsinki. All participants or guardians signed written informed consent for participating in this study.

### Consent to publish

Not applicable.

### Authors’ contributions

G, KI, and NA conceived the study. G drafted the manuscript, and MSH, HW, and TW critically revised the manuscript for important intellectual content. M, DAP, ITa, KAV, MBA, GCG, LSE, BCN, ETG, AAD, AAK, L, and SHI performed the library preparation and NGS. G, MSH, M, VS, Sl, IT, ES, REK, KI, A, S, I, NA, EWD, DAAN, YP, SK, IL, NRA, EA, TN, and TW collected the data; and G, MSH, HW, and M analyzed the data. All authors have read and approved the manuscript and agreed to be accountable for all aspects of the work in ensuring that questions related to the accuracy or integrity of any part of the work are appropriately investigated and resolved.

## Acknowledgments

We thank the Collaborator Members of the Yogyakarta-Central Java COVID-19 study group: Eko Budiono, Heni Retnowulan, Sumardi, Bambang Sigit Riyanto, Munawar Gani, Satria Maulana, Ira Puspitawati, Osman Sianipar (Faculty of Medicine, Public Health and Nursing, Universitas Gadjah Mada [FK-KMK UGM]/RSUP Dr. Sardjito), Bagoes Poermadjaja (Balai Besar Besar Veteriner Wates, Yogyakarta), Indaryati and Havid Setyawan (Balai Besar Teknik Kesehatan Lingkungan dan Pengendalian Penyakit, Yogyakarta), Ludhang Pradipta Rizki and Sri Fatmawati (FK-KMK UGM), Safitriani and Muhammad Taufiq Soekarno (PT. Pandu Biosains). We gratefully acknowledge the authors, the originating and submitting laboratories for their sequence and metadata shared through GISAID. We also thank to Besar Besar Veteriner Wates (Disease Investigation Center Wates) where the virus samples were sequenced using the NGS Illumina MiSeq instrument. All submitters of data may be contacted directly via www.gisaid.org. The Acknowledgments Table for GISAID is reported as Supplementary Table 1.

